# Positive long-term outcomes of functional nasal surgery and histological findings in post-COVID-19 olfactory dysfunction

**DOI:** 10.64898/2026.07.22.26358656

**Authors:** Aagat Sharma Khatiwada, Alfonso Luca Pendolino, Ziyi Liu, Bruno Scarpa, Matthew S Grubb, Peter J Andrews

## Abstract

**Background:** Olfactory dysfunction (OD) is common in COVID-19 and in a small percentage of patients it can become persistent, substantially impacting on quality of life. Effective treatment options for persistent COVID-19-related OD (C19OD) remain limited. Functional septorhinoplasty (fSRP) can improve olfaction in persistent C19OD by increasing nasal airflow to the olfactory cleft and improving olfactory epithelium (OE) stimulation. However, this implies a retained and functioning OE in C19OD patients. This study evaluates the 12-month outcomes of fSRP and investigates the histopathological correlates supporting the role of nasal airflow improvement to restore olfaction.

**Methods:** This observational cohort study enrolled 12 patients with persistent C19OD who underwent fSRP. Olfactory function (Sniffin’ Sticks TDI scores) and nasal airflow were assessed at baseline, 3, 6, and 12 months. Endoscopic olfactory mucosa biopsies were obtained at baseline from 8 patients for immunohistochemical analysis.

**Results:** fSRP led to statistically significant and clinically relevant TDI improvements at 12 months (median improvement 8.0 from baseline, p<0.005) accompanied by sustained improvement of nasal airflow (p=0.01). Histological analysis showed the presence of immature and mature olfactory sensory neurons in 3 patients (37.5%). In one specimen, intact OE was identified containing horizontal basal cells, indicating preserved regenerative capacity more than two years post-infection.

**Conclusions:** fSRP provides sustained clinical benefits for patients with persistent C19OD. The survival of sensory neurons and progenitor cells in the olfactory mucosa beyond two years suggests a biological substrate for olfactory recovery, which may be facilitated by surgically improved nasal airflow and OE stimulation.

## INTRODUCTION

Olfactory dysfunction (OD) is a salient feature following SARS-CoV-2 (COVID-19) infection, occurring in around 48% of infected subjects.^1^ In a significant proportion of patients, OD can persist beyond 3 months, whereby it is classified as persistent COVID-19-related OD (C19OD).^2,3^ The mechanisms behind persistent C19OD are not fully understood. However, in persistent cases, a chronic inflammation at the level of the olfactory mucosa (OM) is thought to be involved leading to a reduction in the density of olfactory sensory neurons (OSNs).^4,5^

Treatment options for persistent C19OD remain limited. Olfactory training (OT) is regarded worldwide as the gold standard treatment for post-infectious OD (PIOD).^6,7^ However, despite its potential therapeutic value, OT efficacy is shown to be limited in roughly 50% to 85% of individuals,^8^ with up to 29% of PIOD cases failing to demonstrate any olfactory improvement even following a prolonged course of OT.^9^ Moreover, evidence suggests that OT is more useful if started within 12 months from PIOD onset^10^ and, therefore, might be less effective in patients with more longstanding OD. In addition to this, patients’ compliance linked to the long treatment duration required (recommended consistently for 3-6 months minimum) remains an often underestimated limitation.^11,12^ No other pharmacologic or non-pharmacologic treatment option has shown consistent benefit in persistent C19OD.

Functional septorhinoplasty (fSRP) is a nasal procedure which typically restores nasal airflow through correcting a deviated nasal septum and augmenting the internal and external nasal valves (INV/ENV). The INV, in particular, is a crucial anatomical region in the regulation of nasal airflow to the olfactory region.^13^ fSRP can therefore play a role in further increasing olfactory cleft nasal airflow through INV augmentation, which is achieved by insertion of spreader grafts.^14,15^ Previous studies have shown that fSRP can improve olfactory function in subjects with longstanding OD of various aetiologies including PIOD and post-traumatic OD.^16–18^ The mechanisms through which fSRP can improve olfaction in persistent C19OD are not fully understood, but emerging evidence points towards mechanisms beyond correction of just an underlying conductive loss. Growing evidence suggests that by increasing the nasal airflow through the olfactory area, fSRP can improve persistent OD through increased delivery of odorants to the olfactory epithelium (OE), leading to a greater stimulation or reactivation of the remaining OSNs.^18,19^

In 2022 we conducted a prospective-controlled study on subjects with persistent C19OD longer than 2 years and showed that fSRP could lead to statistically significant, and clinically relevant, improvement in the olfactory function at 6 months, demonstrating superiority over OT alone.^18^ To further investigate the pathophysiology underpinning persistent C19OD, and to better understand how fSRP could restore olfaction, we also took targeted endoscopic biopsy of the OM in patients undergoing fSRP. In the present study, we present the 12-month follow-up outcomes of the original cohort and report the histological results of the OM biopsies. Based on our findings and histological results, we discuss the possible mechanistic basis through which fSRP can lead to smell improvement in subjects with persistent C19OD.

## MATERIALS AND METHODS

### Participants

This observational cohort study enrolled consecutive patients presenting to the long-COVID smell clinic at University College London Hospitals, London, United Kingdom, between October 2022 and May 2023. Eligibility criteria have been previously reported.^18^ Eligible patients underwent fSRP and were assessed at baseline (T_0_), 3- (T_1_), 6- (T_2_), and 12-month (T_3_) postoperatively.

### Functional septorhinoplasty and olfactory mucosa (OM) biopsy

fSRP involved septoplasty, nasal bone realignment, and INV and ENV augmentation using bilateral autologous spreader grafts and columellar graft. OM was obtained by taking a single endoscopic biopsy of the superior turbinate, as previously described.^20^ All operations were performed by the same team (PJA/ALP) following the same surgical technique.

### Olfactory testing and airflow measurements

Odour threshold (T), discrimination (D), and identification (I) were evaluated using the Sniffin’ Sticks (S’S) extended set (Burghart, Medisense). Normosmia was defined as a TDI score of ≥30.75, hyposmia as TDI between 16 and 30.75, and functional anosmia as TDI ≤16.^21^ The primary outcome was the minimal clinically important difference (MCID), defined as an improvement by 5.5 points for overall TDI, 2.5 points for threshold, and 3 points for both discrimination and identification.^22^

Self-assessment of olfaction was performed using a visual analogue scale for smell (sVAS; 0 represents “sense of smell absent” and 10 “sense of smell not affected”). Qualitative olfactory dysfunction (i.e., parosmia/phantosmia) was investigated by asking the participants if the symptom was present or not at the moment of the examination.

Bilateral and unilateral peak nasal inspiratory flow (PNIF) measurements were done to assess nasal airflow. Acoustic rhinometry (AR) was used to obtain unilateral first minimal cross-sectional area (MCA1) and nasal volume (NV).^23–25^

### Sample collection and immunohistochemistry

Tissues were fixed in 10% neutral buffered formalin and processed into formalin-fixed, paraffin-embedded (FFPE) blocks within 24 hours of collection. Blocks were microtome-sectioned at 8 μm. Sections were slide-mounted and dried overnight at room temperature (RT), before deparaffinisation (xylene 2 x 5 min) and rehydration (in industrial methylated spirit: 100%, 2 min; 90 %, 2 min, 70 %; 2 min; 50 %, 2 min). After rinsing in phosphate-buffered saline (PBS; 2×5 min), slides were immersed in retrieval buffer (0.1 M trisodium citrate + 0.1 M citric acid in dH2O; 95 °C, 30 min), followed by gradual cooling to RT and 3×5 min PBS washes. Sections were permeabilised and blocked (10 % normal goat serum (NGS) and 0.1 % Triton X-100 in PBS; 2 h, RT), before primary antibody incubation (combinations of anti-Krt5 (Abcam, AB52635), anti-OMP (Abcam, 54-1001), anti-Sox2 (Invitrogen, 14-9811-82), anti-Tuj1 (Biolegend, 801202); all 1:1000 in PBS with 10% NGS and 0.1% Triton X-100; 4 °C, overnight. Slides were then washed (PBS, 3×5 min) before incubation with species-specific, fluorophore-conjugated IgG secondary antibodies (Thermo Fisher Scientific; 1:1000 in PBS with 10% NGS and 0.1% Triton; RT, 2 h). After final washes (PBS, 3 x 5 min), slides were coverslipped (Fluosave mounting medium; Merck Millipore).

### Imaging, image analysis and quantification

Images were acquired with a laser scanning confocal microscope (Zeiss LSM710) with a 40× oil immersion objective, a pinhole of 1 AU, and appropriate excitation and emission filters. Z-stacks (step 0.44 μm; 1024 x 1024 pixels) were acquired at 0.415, 0.138 or 0.104 μm/pixel. For cell density calculations, images were automatically stitched (ImageJ),^26^ and the length of the Tuj1-positive cell-containing region was measured along its apical surface. Positively labelled cells were identified using the Cell Counter plugin based on distinct fluorescent signal above background. Co-localisation for double-labelled cells was confirmed by visual inspection of overlaid channels.

### Statistical Analysis

Quantitative variables were summarized using median and interquartile range whereas qualitative variables were described with frequency and percentage. Comparisons of measurements between baseline and follow-ups were performed using the Mann-Whitney test for quantitative variables and the proportion test for dichotomic variables. Pearson correlation index was used to measure associations between quantitative variables. *p*-values were calculated for all tests, and 5% was considered as the critical level of significance.

## RESULTS

Twelve patients underwent fSRP but only 8 completed the 12-month follow-up (T_3_). Baseline population characteristics are summarised in *Table 1*. At baseline (T_0_), all patients showed impaired olfactory function (S’S threshold and identification below age-adjusted norms), and reduced nasal airflow compared with population normality scores.^25,27–29^ At T_2_ and T_3_ patients demonstrated statistically significant improvements in S’S TDI scores and all subdomains, with changes exceeding the MCID for threshold and identification at T_2_ and both TDI and all subdomains at T_3_. (*Figure 1; Table 2–3*) sVAS significantly improved at T_3_ (p=0.04). At T_3_, 2 (2/8, 25%) patients had parosmia, and one (1/8, 13%) had phantosmia. This compares with 9/11 patients (81%) who had parosmia and 3/11 (27%) with phantosmia at baseline, and 4/9 (44%) with parosmia and 0/9 (0%) with phantosmia at 6 months. At T_2_ only the bilateral and the right nasal volume significantly improved (p<0.05) whiles at T_3_ both the bilateral and unilateral PNIF, as well as the nasal volumes, significantly improved (p<0.05). *(Table 3)*

**Figure 1.**
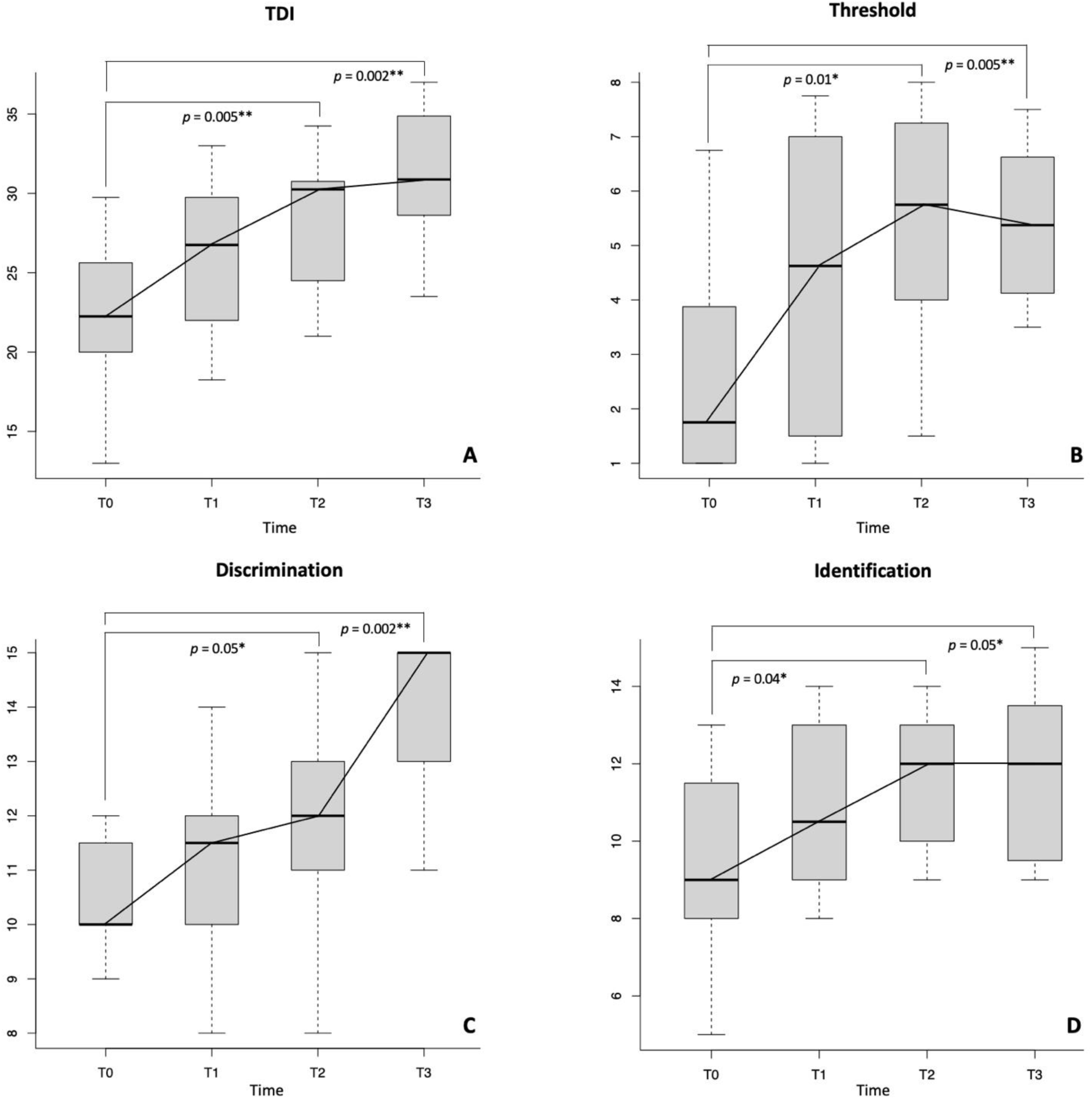
Box plots showing changes in TDI (a), threshold (b), discrimination (c) and identification (d) scores for the fSRP group during the study period. Statistical difference between T_0_-T_2_ and T_0_-T_3_ is also shown. Levels of significance *p ≤ 0.05, **p ≤ 0.01. TDI: Threshold + Discrimination + Identification.

**Table 1.** General characteristics of the population at baseline. +Length of OD is calculated as number of days from the infection date to the day of enrolment. OD: olfactory dysfunction; PIOD: post-infectious olfactory dysfunction.

|  | *n* = 12 |
| --- | --- |
| Age, median [P25-P75], yr | 40.0 [31.5-44.0] |
| Sex, No (%) |  |
| Female | 9 (75.0%) |
| Male | 3 (25.0%) |
| Length of OD<sup>+</sup>, median [P25-P75], yr | 2.3 [2.0-2.5] |
| Parosmia, No (%) | 10 (83.3%) |
| Phantosmia, No (%) | 4 (33.3%) |
| Smoking, No (%) |  |
| Ex-smoker | 1 (100%) |
| Yes | 0 (0.0%) |
| No | 0 (0.0%) |
| Comorbidity, No (%) |  |
| None | 8 (66.7%) |
| Yes | 4 (33.3%) |
| Hypothyroidism | 1 (25.0%) |
| Asthma | 1 (25.0%) |
| Others | 4 (100%) |
| Allergic rhinitis, No (%) | 2 (16.7%) |
| Chronic rhinosinusitis, No (%) | 0 (0.0%) |
| Family history Alzheimer/Parkinson, No (%) | 3 (25.0%) |
| History of PIOD, No (%) | 2 (16.6%) |
| History of previous nasal operations, No (%) | 0 (0.0%) |
| History of head trauma, No (%) | 1 (8.3%) |

**Table 2.** Olfactory and nasal measurements, and patient-reported outcome measures (PROMs) at baseline, 3, 6, and 12-month following functional septorhinoplasty. TDI: Threshold + Discrimination + Identification; PNIF; peak nasal inspiratory flow; lPNIF: left PNIF; rPNIF: right PNIF; MCA1: first minimal cross-sectional area. SF-36: 36-item Short Form Survey; sVAS: Visual Analogue Scale for sense of smell; SNOT-22: 22-item SinoNasal Outcome Test; NOSE: Nasal Obstruction and Septoplasty Effectiveness Scale

|  | Baseline (T<sub>0</sub>)<br>n=12 | 3-month (T<sub>1</sub>)<br>n=10 | 6-month (T<sub>2</sub>)<br>n=9 | 12-month (T<sub>3</sub>)<br>n=8 |
| --- | --- | --- | --- | --- |
| **Sniffin' Sticks** |  |  |  |  |
| TDI, median [P25-P75] | 22.3 [20.0-24.8] | 26.8 [20.0-24.8] | 30.3 [24.5-30.8] | 30.3 [28.5-33.3] |
| Threshold, median [P25-P75] | 1.8 [1.0-3.8] | 4.6 [1.7-7.0] | 5.8 [4.0-7.3] | 5.4 [4.3-6.4] |
| Discrimination, median [P25-P75] | 10.0 [10.0-11.3] | 11.5 [10.0-12.0] | 12.0 [11.0-13.0] | 15.0 [14.0-15.0] |
| Identification, median [P25-P75] | 9.0 [8.0-11.3] | 10.5 [9.0-12.8] | 12.0 [10.0-13.0] | 12.0 [9.8-13.3] |
| Normosmics, n (%) | 0 (0.0%) | 1 (10.0%) | 4 (44.4%) | 5 (62.5%) |
| Hyposmics, n (%) | 11 (91.7%) | 9 (90.0%) | 5 (55.6%) | 3 (37.5%) |
| Anosmics, n (%) | 1 (8.3%) | 0 (23.5%) | 0 (0.0%) | 0 (0.0%) |
| **Nasal measurements** |  |  |  |  |
| PNIF, median [P25-P75], L/min |  |  |  |  |
| Bilateral PNIF | 115.0 [87.5-137.5] | 137.5 [130.0-157.5] | 160.0 [125.0-190.0] | 170.0 [140.0-202.5] |
| Right PNIF | 62.5 [50.0-82.5] | 82.5 [66.3-130.0] | 110.0 [70.0-120.0] | 107.5 [83.8-120.0] |
| Left PNIF | 60.0 [48.8-77.5] | 90.0 [81.3-107.5] | 65.0 [50.0-100.0] | 105.0 [83.8-117.5] |
| Acoustic rhinometry, median [P25-P75] |  |  |  |  |
| Right MCA1, cm<sup>2</sup> | 0.5 [0.4-0.7] | 0.5 [0.4-0.7] | 0.6 [0.4-0.7] | 0.8 [0.7-0.9] |
| Right Nasal volume (0-5), cm<sup>3</sup> | 5.7 [5.2-7.3] | 8.1 [5.9-10.9] | 7.9 [5.8-9.4] | 10.0 [7.8-14.2] |
| Left MCA1, cm<sup>2</sup> | 0.7 [0.4-0.8] | 0.7 [0.5-0.8] | 0.6 [0.5-0.8] | 0.8 [0.6-0.9] |
| Left Nasal volume (0-5), cm<sup>3</sup> | 6.6 [5.7-7.4] | 9.5 [7.6-10.7] | 8.0 [6.0-9.1] | 12.3 [9.5-15.7] |
| **PROMs** |  |  |  |  |
| SF-36, median [P25-P75], % |  |  |  |  |
| Physical functioning | 95.0 [83.8-100] | 100 [81.3-100] | 95.0 [90.0-100] | 92.5 [80.0-100] |
| Role limitations due to physical health | 62.5 [25.0-100] | 75.0 [31.3-100] | 75.0 [50.0-100] | 100.0 [18.8-100] |
| Role limitations due to emotional problems | 100 [33.3-100] | 100 [66.7-100] | 100 [100-100] | 100 [91.7-100] |
| Energy/Fatigue | 50.0 [23.8-73.8] | 60.0 [37.5-75.0] | 55.0 [50.0-80.0] | 57.5 [36.3-82.5] |
| Emotional wellbeing | 70.0 [67.0-88.0] | 72.0 [68.0-88.0] | 84.0 [76.0-88.0] | 86.0 [75.0-93.0] |
| Social functioning | 68.8 [46.9-100] | 100 [65.6-100] | 100 [81.3-100] | 93.8 [46.9-100] |
| Pain | 78.8 [65.0-90.0] | 90.0 [78.1-97.5] | 90.0 [73.8-95.0] | 80.0 [64.4-92.5] |
| General health | 65.0 [45.0-72.5] | 62.5 [55.0-83.8] | 75.0 [50.0-85.0] | 67.5 [36.3-83.8] |
| sVAS, median [P25-P75] | 4.3 [3.0-5.3] | 5.0 [4.0-6.4] | 6.0 [4.5-6.0] | 7.0 [5.4-7.3] |
| SNOT-22, median [P25-P75] | 25.0 [14.3-30.0] | 13.0 [9.3-32.5] | 11.0 [6.0-15.0] | 17.5 [9.8-31.5] |
| NOSE, median [P25-P75] | 25.0 [12.5-45.0] | 17.5 [7.5-34] | 10.0 [10.0-15.0] | 20.0 [5.0-31.5] |

**Table 3.** Differences in medians and statistical significance (p-values – in brackets). The sign ‘+’ indicates an improvement while the sign ‘-‘ indicates a worsening in the median values. Significant p-values in bold. Levels of significance *p ≤ 0.05, **p ≤ 0.01. TDI: Threshold + Discrimination + Identification; PNIF; peak nasal inspiratory flow; lPNIF: left PNIF; rPNIF: right PNIF; MCA1: first minimal cross-sectional area. PROMs: patient-reported outcome measures; SF-36: 36-item Short Form Survey; sVAS: Visual Analogue Scale for sense of smell; SNOT-22: 22-item SinoNasal Outcome Test; NOSE: Nasal Obstruction and Septoplasty Effectiveness Scale

|  | T<sub>0</sub>-T<sub>1</sub> | T<sub>1</sub>-T<sub>2</sub> | T<sub>2</sub>-T<sub>3</sub> | T<sub>0</sub>-T<sub>2</sub> | T<sub>0</sub>-T<sub>3</sub> |
| --- | --- | --- | --- | --- | --- |
| **Sniffin' Sticks** |  |  |  |  |  |
| TDI | +4.5 (0.15) | +3.5 (0.22) | +0.0 (0.50) | +8.0 (**0.005**)<sup>**</sup> | +8.0 (**0.002**)<sup>**</sup> |
| Threshold | +2.8 (0.08) | +1.2 (0.57) | -0.4 (0.96) | +4.0 (**0.01**)<sup>**</sup> | +3.6 (**0.005**)<sup>**</sup> |
| Discrimination | +1.5 (0.46) | +0.5 (0.28) | +3.0 (0.06) | +2.0 (**0.05**)<sup>*</sup> | +5.0 (**0.002**)<sup>**</sup> |
| Identification | +1.5 (0.27) | +1.5 (0.38) | +0.0 (0.92) | +3.0 (**0.04**)<sup>*</sup> | +3.0 (**0.05**)<sup>*</sup> |
| **Nasal measurements** |  |  |  |  |  |
| PNIF, L/min |  |  |  |  |  |
| Bilateral PNIF | +22.5 (0.06) | +22.5 (0.84) | +10.0 (0.44) | +45.0 (**0.04**)<sup>*</sup> | +55.0 (**0.01**)<sup>**</sup> |
| Right PNIF | +20.0 (0.11) | +27.5 (0.77) | -2.5 (0.70) | +47.5 (0.07) | +45.0 (**0.005**)<sup>**</sup> |
| Left PNIF | +30.0 (**0.03**)<sup>*</sup> | -25.0 (0.27) | +40.0 (0.18) | +5.0 (0.40) | +45.0 (**0.03**)<sup>*</sup> |
| Acoustic rhinometry |  |  |  |  |  |
| Right MCA1, cm<sup>2</sup> | 0.0 (0.62) | +0.1 (0.29) | +0.2 (0.25) | +0.1 (0.34) | +0.3 (0.09) |
| Right Nasal volume (0-5), cm<sup>3</sup> | +2.4 (0.07) | -0.2 (0.66) | +2.1 (0.67) | +2.2 (**0.03**)<sup>*</sup> | +4.3 (**0.01**)<sup>**</sup> |
| Left MCA1, cm<sup>2</sup> | 0.0 (0.72) | -0.1 (0.96) | +0.2 (0.78) | -0.1 (0.84) | +0.1 (0.41) |
| Left Nasal volume (0-5), cm<sup>3</sup> | +2.9 (0.09) | -1.5 (0.90) | +4.3 (0.28) | +1.4 (0.11) | +5.7 (**0.02**)<sup>*</sup> |
| **PROMs** |  |  |  |  |  |
| SF-36, % |  |  |  |  |  |
| Physical functioning | +5.0 (0.55) | -5.0 (0.83) | -2.5 (0.62) | 0.0 (0.74) | -2.5 (0.87) |
| Role limitations due to physical health | +10.0 (0.68) | 0.0 (0.97) | +25.0 (0.79) | +10.0 (0.61) | +37.5 (0.57) |
| Role limitations due to emotional problems | 0.0 (0.85) | 0.0 (0.40) | 0.0 (0.95) | 0.0 (0.47) | 0.0 (0.57) |
| Energy/Fatigue | +10.0 (0.69) | -5.0 (0.62) | +2.5 (0.81) | +5.0 (0.41) | +7.5 (0.70) |
| Emotional wellbeing | +2.0 (0.74) | +12.0 (0.44) | +2.0 (0.81) | +14.0 (0.43) | +16.0 (0.20) |
| Social functioning | +31.2 (0.24) | 0.0 (0.93) | -6.2 (0.57) | +31.2 (0.31) | +25.0 (0.84) |
| Pain | +11.2 (0.57) | 0.0 (0.65) | -10.0 (0.92) | +11.2 (0.74) | +2.2 (0.94) |
| General health | -2.5 (0.74) | +12.5 (0.97) | -7.5 (1) | +10.0 (0.50) | +2.5 (0.97) |
| sVAS | +0.7 (0.34) | +1.0 (0.84) | +1.0 (0.17) | +1.7 (0.17) | +2.7 (**0.04**)<sup>*</sup> |
| SNOT-22 | -12.0 (0.27) | -2.0 (0.35) | +6.5 (0.25) | -14.0 (**0.03**)<sup>*</sup> | -7.5 (0.61) |
| NOSE | -7.5 (0.48) | -7.5 (0.20) | +10 (0.30) | -15.0 (**0.05**)<sup>*</sup> | -5.0 (0.43) |

### Correlation Between Olfactory Function and Nasal Measurements

When we looked at the correlations between the changes in S’S scores and nasal measurements between T_0_ and T_2_, we found strong significant correlations between changes in left PNIF and changes in TDI (r=0.67; p=0.05), between changes in total PNIF and changes in discrimination (r=0.73; p=0.03) and identification (r=0.67; p=0.05), and between changes in left MCA1 and changes in identification (r=0.74; p=0.03). Between T_0_ and T_3_ strong significant correlations were observed between changes in identification and changes in right/left MCA (r=0.77; p=0.03; r=0.83; p=0.02) and left NV (r=0.75; p=0.03).

### Sensory axon bundles in cases of long-term C19OD

OM biopsies from 8 patients underwent immunohistochemical staining and were examined for the presence of neural tissue. In three cases (samples 2, 4 and 5) the neuronal marker Tuj1^30^ labelled clear bundles of OSN axons. (*Figure 2*) A minority of axons within each of these bundles were also positive for the selective marker of mature OSNs, olfactory marker protein (OMP; *Figure 2*).^30^ The presence of these axon bundles strongly suggests the retained integrity of some immature (Tuj1-positive only) and mature (Tuj1- and OMP-positive) OSNs in these three cases.

**Figure 2.**
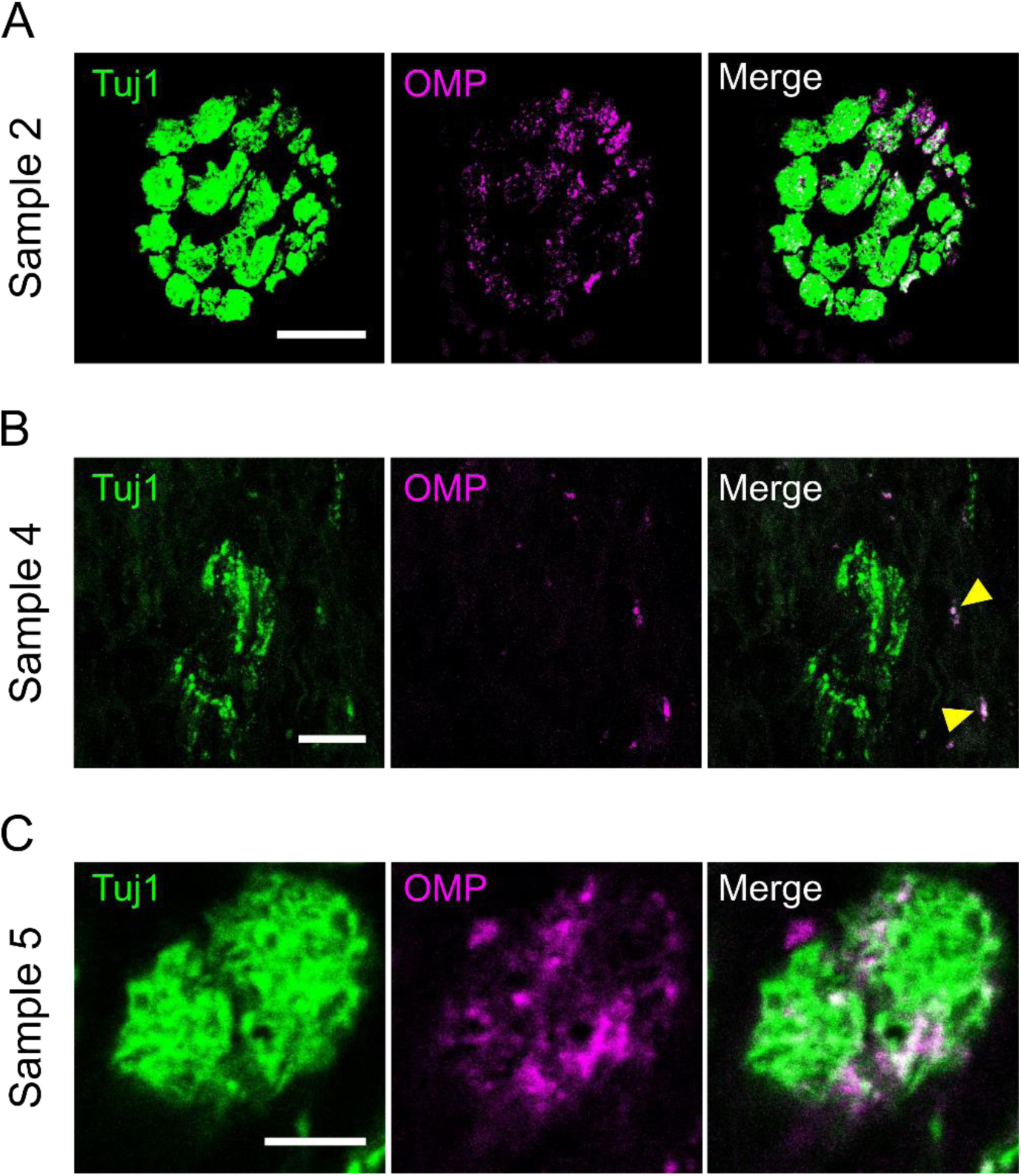
Olfactory sensory neuron axon bundles in cases of long-term post-COVID olfactory dysfunction. **A)** Example single-plane confocal image of a sensory neuron axon bundle in biopsy sample 2, immunolabelled for Tuj1 (all olfactory sensory neurons) and OMP (mature olfactory sensory neurons). Scalebar, 20 µm. **B)** Example maximum intensity projection confocal image of sensory neuron axons in biopsy sample 2, labelled for Tuj1 and OMP. Scalebar, 10 µm; yellow arrowheads, isolated double-labelled axons. **C)** Example single-plane confocal image of a small sensory neuron axon bundle in biopsy sample 5, labelled for Tuj1 and OMP. Scalebar, 5 µm.

### Olfactory epithelial composition in a case of long-term C19OD

In one nasal biopsy sample (sample 5) we also observed a region of tissue with all the cellular hallmarks of the OE. Although uneven with signs of some physical damage (*Figure 3A*), this region was clearly demarcated by Tuj1-positive OSNs^4^ (*Figure 3A,B*; 0.22 cells/µm). Krt5-positive horizontal basal cells (HBC) were also present, in a multicellular basal layer^31^ (*Figure 3A,B*; 0.13 cells/µm). Labelling for Sox2 revealed positive nuclei in an apical layer of sustentacular cells, and in a layer of basal cells (*Figure 3A,B*; 0.07 and 0.09 cells/µm).^4,30,31^ In the basal OE, co-label of Krt5 and Sox2 was observed in 50 % of all Krt5-positive cells (62/125), and in 76 % of all Sox2-positive cells (62/82; *Figure 3C*). When we stained for OMP, we observed mature OSNs that all co-labelled with Tuj1^30,32^ (*Figure 3D-F*; 0.05 cells/µm), but that comprised only 22 % (47/210) of all OSNs in this sample. The ratio between OMP-positive neurons and apical Sox2-positive sustentacular cells^4^ was 0.34 (47/140).

**Figure 3.**
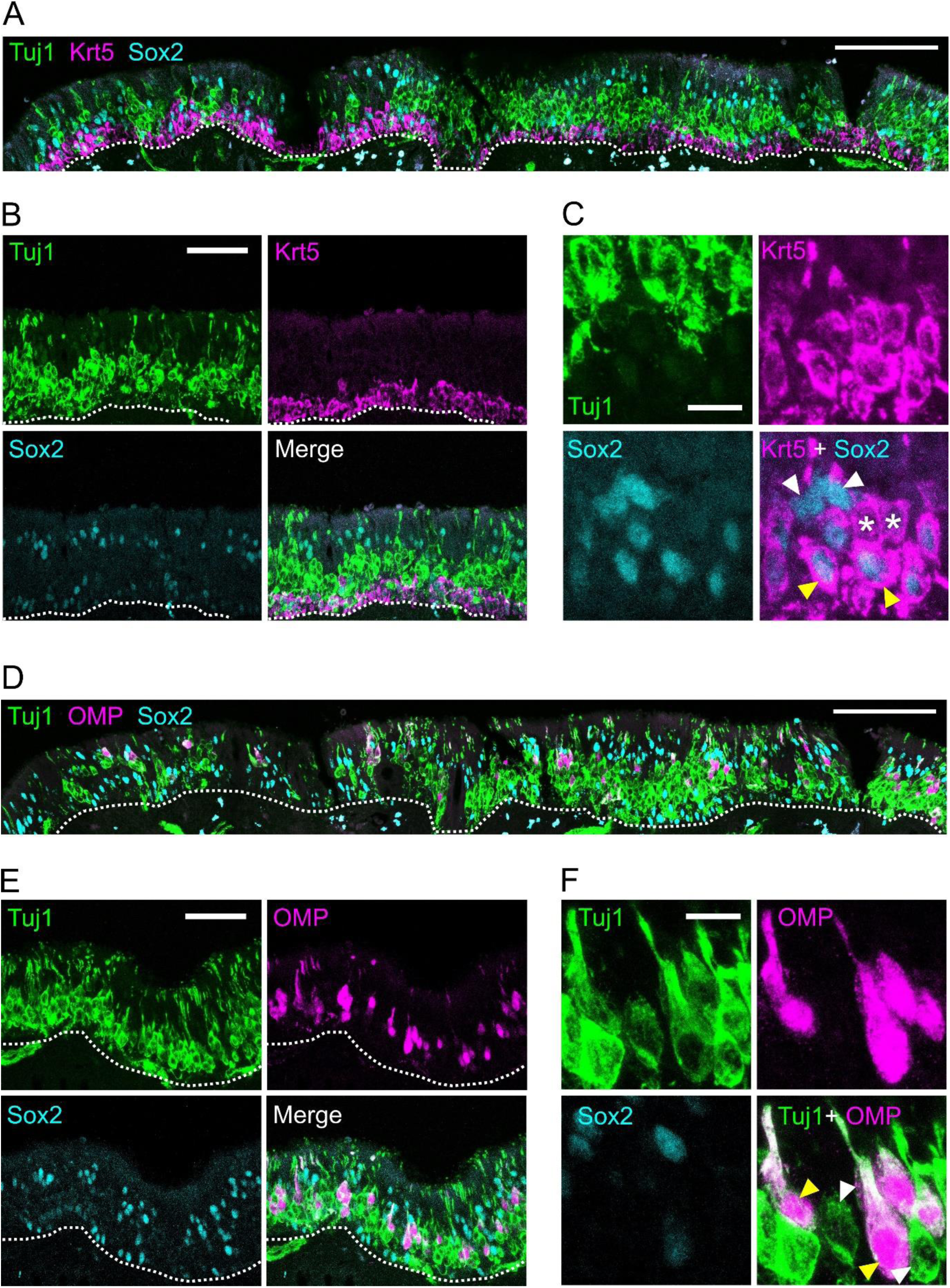
Olfactory epithelial composition **(A-C)** and low density of mature olfactory sensory neurons **(D-F)** in a case of long-term post-COVID olfactory dysfunction. **A)** Example maximum intensity projection stitched confocal image of olfactory epithelium in biopsy sample 5, immunolabelled for Tuj1 (olfactory sensory neurons), Krt5 (horizontal basal cells), and Sox2 (sustentacular and basal cells). Scalebar, 100 µm; dotted line, basal boundary of olfactory epithelium. **B)** Example maximum intensity projection confocal images of Tuj1, Krt5, Sox2 and merged label in a region of olfactory epithelium in biopsy sample 5. Scalebar, 50 µm; dotted line, basal boundary of olfactory epithelium. **C)** Example cellular-level maximum intensity projection confocal image of Tuj1, Krt5, Sox2 and merged label in a basal portion of olfactory epithelium in biopsy sample 5. Scalebar, 10 µm; asterisks, cells positive for Krt5 only; white arrowheads, cells positive for Sox2 only; yellow arrowheads, cells positive for both Krt5 and Sox2. **D)** Example maximum intensity projection stitched confocal image of olfactory epithelium in biopsy sample 5, immunolabelled for Tuj1 (all olfactory sensory neurons), OMP (mature olfactory sensory neurons), and Sox2 (sustentacular and basal cells). Scalebar, 100 µm; dotted line, basal boundary of olfactory epithelium. **E)** Example maximum intensity projection confocal images of Tuj1, OMP, Sox2 and merged label in a region of olfactory epithelium in biopsy sample 5. Scalebar, 50 µm; dotted line, basal boundary of olfactory epithelium. **F)** Example cellular-level maximum intensity projection confocal image of Tuj1, OMP, Sox2 and merged label in a portion of olfactory epithelium in biopsy sample 5. Scalebar, 10 µm; white arrowheads, cells positive for Tuj1 only; yellow arrowheads, cells positive for both Tuj1 and OMP.

## DISCUSSION

Our longitudinal study confirmed a sustained improvement in olfactory function after 12 months following fSRP. In fact, at both T_2_ and T_3_, the overall TDI score, as well as all subdomains of the S’S olfactory test showed statistically significant improvement with MCID reached at T_3_ for all subdomains. At 12 months, the largest contribution in the TDI improvement was driven by the discrimination subdomain, while improvement in threshold drove the largest improvement at 6 months (*Table 2–3*). Odour threshold is thought to be more sensitive to peripheral olfactory circuit changes, whereas changes in odour discrimination are believed to be driven more by alterations in central olfactory circuits.^33,34^ This may suggest that fSRP initially leads to improved activity at an OE level, while in the longer term it potentiates plasticity of higher olfactory centres.

The olfactory improvement was accompanied by, and likely driven by, increased PNIF values both unilaterally and bilaterally, with an average increase of 40% at T_2_ and 48% at T_3_ (*Table 2–3*). Amongst the available measures of nasal airflow, PNIF may best reflect olfactory airflow, as it is assessed during forced sniffing, a manoeuvre commonly used to enhance odour perception.^35^

Histological analysis of the OE in our cohort at baseline provides potential mechanistic insight into the cellular basis of persistent C19OD, and offers further clues as to how fSRP can improve olfactory ability. We found evidence for retention of OSNs in 3 out of 8 subjects who had persistent C19OD lasting at least 2 years. Moreover, in one of our histology samples, we were also able to clearly identify all major cellular components and intact laminar structure of the OE. This included a significant population of OSNs, as well as a dense band of HBCs, OE progenitors which become activated in situations of significant OE injury but remain otherwise dormant.^36,37^ Despite the reported downregulation of genes involved in epithelial renewal in the HBCs of patients with persistent C19OD,^4^ the significant population of HBCs present in our biopsy sample suggests that a reserve of progenitor cells capable of at least partially regenerating the OE remains available even beyond 2 years after SARS-CoV-2 infection. Together, these observations imply the possible retention of some olfactory ability even after years of persistent C19OD, and are entirely consistent with the hyposmic, rather than fully anosmic status of our cohort prior to surgery.

However, we also observed a stark reduction in mature OSNs (mOSNs) in our OE-containing biopsy, consistent with the olfactory deficits recorded in our patients at pre-surgical baseline. A previous study found a marked reduction in the ratio of mOSNs to sustentacular cells of ∼0.1 in persistent C19OD, compared with ∼0.75 in normosmic patients.^4^ In our sample, we found a ratio of 0.34, which is approximately halfway between these values. This may be explained by a gradual spontaneous recovery of OSNs that occurs over time in persistent C19OD, since Finlay et al.^4^ included participants who had persistent C19OD lasting around 3 months compared to our cohort of subjects who had persistent C19OD lasting at least 2 years.

Another interesting finding in our histological sample was the proportion of immature OSNs (iOSNs). We found this to be 78%, much higher than the proportion reported in healthy olfactory tissue (55%).^31^ This suggests that perhaps there is a deficit of OSN maturation, or a specific susceptibility to cell death in mOSNs in persistent C19OD. Chronic inflammatory changes such as lymphocytic infiltration, and imbalanced macrophage differentiation, have been reported in persistent C19OD, which may impair normal OSN maturation and/or maintenance.^4,5,38^

In all of our 3 cases where we found evidence for the continued presence of sensory neurones, the pre-operative olfactory function was in the hyposmic range (*Figure 2, 5A*). Although the one patient whose biopsy contained a clear section of OE (Sample 5; *Figure 3*) had the highest pre-operative TDI score, the other two patients with OSN axonal label (Samples 2 and 4; *Figure 2*) were in the mid-range of all patient values at this timepoint. After surgery, all 3 patients with OSN-positive biopsies attained TDI improvements after 12 months that exceeded the MCID (*Figure 4B*). However, these were not the three highest overall TDI improvements, and the MCID was also reached in all but one of the patients whose samples were negative for neuronal markers (*Figure 4B*). Moreover, the one patient whose biopsy contained clear evidence for a retained section of OE (Sample 5) achieved only the 5^th^-largest TDI change (*Figure 4B*). These data suggest that, while the presence of any neural tissue in biopsy samples might predict a favourable, clinically important change in olfactory ability, the absence of such histological features does not preclude equally important improvements from being observed.

**Figure 4.**
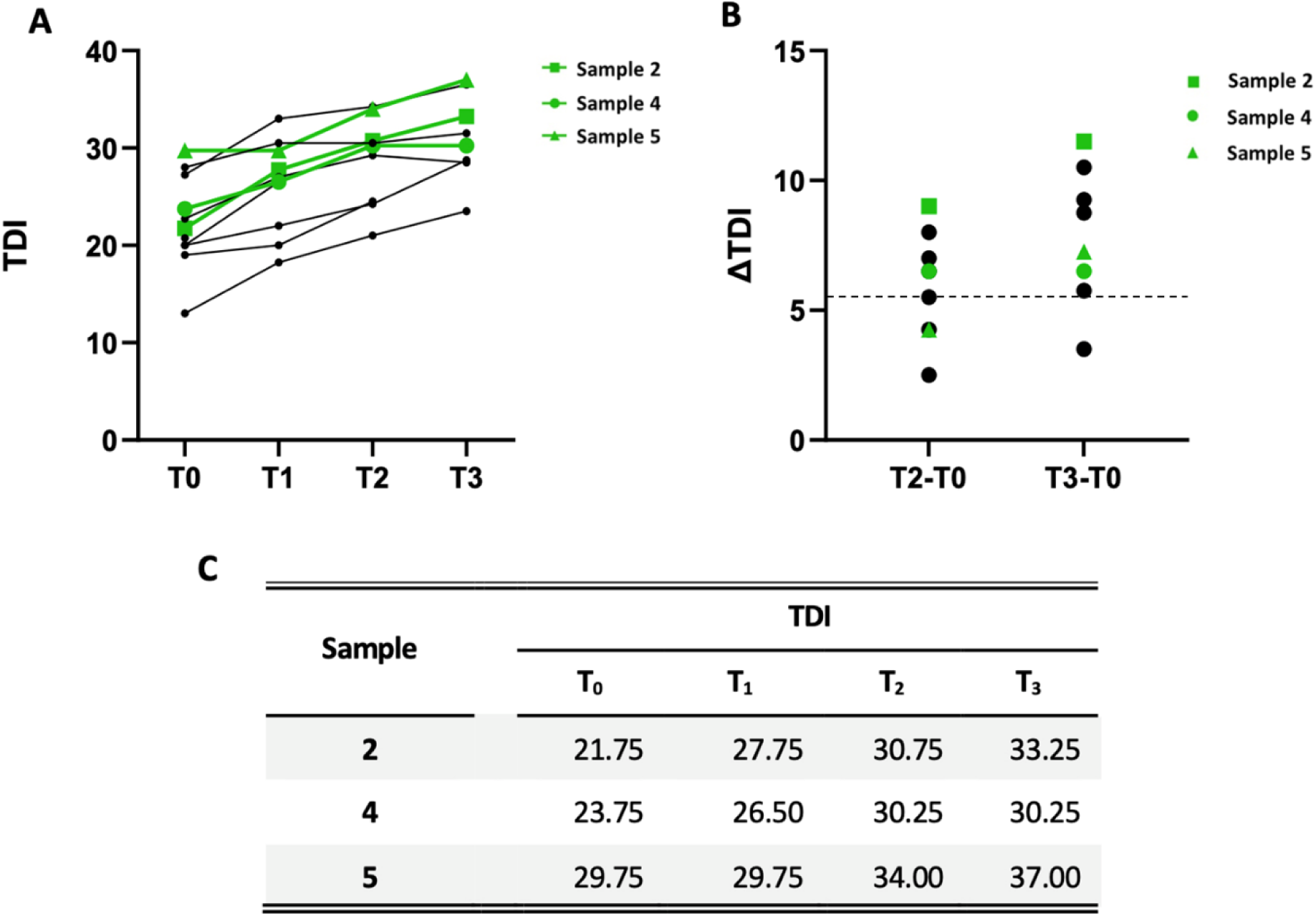
Sensory neuron features in biopsy tissue are not necessarily predictive of olfactory recovery. A) Total Sniffin Sticks score (TDI) at different timepoints (T). Each dot shows TDI for one patient at a given timepoint; lines connect different timepoints of assessment for the same patient; green colouring shows data from patients whose biopsies were confirmed to contain neural tissue. B) Difference in total TDI score between timepoint T0 and T2, and T0 and T3. Each dot represents one patient; green symbols show patients whose biopsies were confirmed to contain neural tissue. Dotted horizontal line represents the MCID threshold. C) Table showing Sniffin Sticks TDI scores for the 3 patients whose biopsies were confirmed to contain neural tissue.

These clinical and histological findings suggest potential mechanisms by which fSRP may improve olfaction in persistent C19OD. The most straightforward explanation is that enhanced nasal airflow increases odorant delivery to the OE, thereby stimulating the remaining OSNs without altering the cellular composition of the OM. In particular, increased airflow within the olfactory cleft achieved through INV augmentation with spreader grafts appears to play a key role. Supporting this hypothesis, our recent prospective-controlled study demonstrated superior outcomes with fSRP compared with OT, with significant improvements in S’S scores observed only in the fSRP group.^18^ This finding suggests that increasing sensory exposure through enhanced airflow beyond what is afforded by OT alone, may be necessary to promote further olfactory recovery in patients with persistent C19OD lasting more than two years. Consistent with this interpretation, changes in PNIF and AR were positively correlated with changes in olfactory scores at 6 and 12 months. These results also align with previous studies reporting associations between improvements in PNIF and odour threshold scores in patients with PIOD.^16,19^

Improved airflow to the OE and its associated increase in sensory stimulation (above that provided by OT) could also promote OSN survival and/or maturation. Indeed, sensory stimulation is important for the survival of OSNs in the OE,^39–42^ and factors which reduce neuronal excitability of OSNs promote their loss.^41–43^ This activity dependent survival and functioning of OSNs may be a key underlying principle in olfactory improvement after fSRP. Given recent reports that even immature OSNs can carry significant odour information in the mouse,^44,45^ an activity-dependent trophic effect on the OE would not necessarily be contradicted by a strong preponderance of immature OSNs in persistent C19OD as seen in our biopsy sample.

On the other hand, it is also possible that improved airflow to the olfactory cleft kickstarts the normal maturation pathway of OSNs. It would be interesting and hugely informative to study the histological changes in respect to the overall OSN density as well as the proportion of iOSNs in the OE of subjects after fSRP, although unfortunately this is not possible due to obvious ethical implications. Equally, moving forwards our findings show that INV augmentation alone could be routinely performed on patients with otherwise normal nasal airflow to help restore persistent post COVID olfactory dysfunction.

### Strengths and limitations

This is the first study reporting the long-term effects of fSRP on persistent C19OD. Our histological findings give meaningful insights into the potential mechanisms through which fSRP improves olfaction. The study adds to the very limited available literature on human OM biopsy in persistent C19OD, and is the first to investigate the OM histology at 2-year post-infection. To this end, our findings shed light on the underlying pathophysiology of persistent C19OD. The main limitations of our study are the small sample size and the drop-out rate through the study. The lack of OE captured in a significant proportion of our histological samples, limiting the interpretation of our findings, could be potentially explained by fibrosis, contraction, retraction or respiratory metaplasia^4,36^ of the OE in persistent C19OD.

## CONCLUSIONS

Our study shows that fSRP can lead to significant and clinically important sustained olfactory improvement in persistent C19OD lasting over 2 years. Histological analysis of OM of persistent C19OD patients shows the persistence of immature OSNs, mature OSNs as well as progenitor cells. However, the proportion of mature OSNs compared to sustentacular cells is reduced, and a high proportion of OSNs remain immature. The increased airflow to the olfactory cleft following fSRP increases odorant delivery, may lead to a trophic response in OSNs, or may stimulate the recovery of the OE by promoting maturation of immature OSNs. The precise mechanistic effect of fSRP on olfactory function and on the OE should be further investigated in larger studies.

## Data Availability

All data produced in the present study are available upon reasonable request to the authors

## ACKNOWLEDGEMENTS

We would like to thank Abigail Tucker, Neal Anthwal and Wai Lin Tsang for histology training and assistance, and members of the Grubb lab for discussion.

## Notes

**Conflict of Interest:** The authors declare that they have no conflict of interest.

### Competing Interest Statement

The authors have declared no competing interest.

### Clinical Trial

NA

### Clinical Protocols

https://www.thieme-connect.de/products/ejournals/abstract/10.1055/a-2535-0153

### Author Declarations

Hospital Research Ethics Committee of University College London Hospitals gave ethical approval for this work (ref: 14/SC/1180)

